# Revisiting Fever of Unknown Origin (FUO): A Single Tertiary Care Center Experience in North India Calls for Criteria Revision

**DOI:** 10.1101/2023.08.03.23293578

**Authors:** Pathik Dhangar, Prasan Kumar Panda, Rajvi Chaudhary, Ravi Kant, Rohit Gupta, Ruchi Dua, Ashutosh Tiwari, Sandeep Saini, Kavita Khoiwal

## Abstract

**Background:** Fever of Unknown Origin (FUO) is a challenging medical condition characterized by prolonged fever without an identifiable cause despite extensive evaluation. The existing definition, proposed in 1961, requires illness lasting over three weeks, fever of ≥38.3°C (≥101°F) on two occasions, and uncertain diagnosis after one week of inpatient evaluation, excluding immunocompromised patients and in-hospital evaluation. However, the current criteria may not fully capture the diverse spectrum of FUO cases, especially in resource-constrained settings like North India. This study aims to revisit the criteria for FUO in terms of durations of non-diagnosis, temperature ranges, and duration of hospitalization rather than considering the duration of illness and using a set of obligatory investigations. It will propose a more practical and effective approach based on local healthcare resources.

**Methods:** This study utilized a retrospective and prospective longitudinal-exploratory design and was conducted at a single tertiary care center, All India Institute of Medical Sciences (AIIMS) in Rishikesh, North India. The study population consisted of 228 patients meeting the inclusion criteria from January 2018 to December 2022. Inclusion criteria involved adult patients with documented fever of ≥99.1°F on at least two occasions and fever lasting for more than three days. Exclusion criteria included patients with a definitive diagnosis within three days of hospitalization. Data were collected from the hospital’s MRD section retrospectively and through prospective follow-up of eligible patients. Proposed new definitions for FUO were considered, encompassing different ranges of duration of non-diagnosis (3-21 days, >21 days) temperature ranges (99.1°F-100.9°F and ≥101°F), and durations of hospitalization (3-7 days and >7 days). The frequency of each definition was measured. The etiology and outcomes of patients under each definition were analyzed using appropriate statistical tests.

**Findings:** Among the proposed definitions, Definition B (fever lasting 3-7 days with temperatures between 99.1°F-100.9°F) had the highest prevalence (40.8%), followed by Definitions A, D, and C. In contrast, only 5 patients (2.2%) met the classical Definition H (in terms of temperature and duration of hospitalization). By closely observing the patients in Definition B group, around 36.5% patient remained non-diagnosed between >7 to 10 days which is highest among the group. Approximately 62% of patients remained undiagnosed before 21 days of hospitalization and initiated treatment, with temperatures ranging between 99.1°F to 100.9°F. The remaining 38% of patients, with temperatures ≥101°F, also remained undiagnosed before commencing treatment. The majority of patients (94%) underwent diagnostic workups and treatment within 21 days of hospitalization, reflecting the importance of early evaluation and intervention.

**Discussion:** The findings of this study underscore the need for revising the criteria for diagnosing FUO. The prevalence of FUO cases under the proposed definitions suggests that the classical criteria may not fully capture the diverse patient population’s needs and local healthcare resources. The study highlights the importance of a flexible diagnostic timeline, comprehensive clinical assessment, and individualized temperature thresholds to ensure timely and effective care for patients with undiagnosed fever. The lack of significant differences in etiology and outcomes among the various definitions further supports the idea that modifying the criteria can enhance the management of FUO cases without compromising patient outcomes.

**Conclusion:** The study emphasizes the need for a revised approach to defining FUO, taking into account the local healthcare resources and patient population. Adapting the criteria based on temperature ranges and durations can lead to a more practical and effective diagnosis and management of FUO. The proposed flexible diagnostic timeline and comprehensive clinical assessment can improve the timely identification of underlying causes and guide clinicians in providing appropriate treatment and support for patients with undiagnosed fever. Continued research and collaboration within the medical community are essential to develop context-specific guidelines for diagnosing and managing FUO.

## INTRODUCTION

Fever of Unknown Origin (FUO) is a condition where the cause of prolonged fever remains unclear despite extensive evaluation. The traditional definition, proposed in 1961, involved illness lasting over three weeks, fever of ≥38.3°C (≥101°F) on two occasions, and uncertain diagnosis after one week of inpatient evaluation. The current definition excludes in-hospital evaluation and immunocompromised patients.

The current criteria for FUO include fever on two occasions with illness lasting three weeks or longer, absence of the immunocompromised state, and uncertain diagnosis despite thorough investigations. Adapting the definition based on local healthcare resources allows for a more practical and effective diagnosis and management of FUO cases.

Managing FUO involves addressing the underlying cause and providing appropriate treatment. If a definitive diagnosis remains elusive despite thorough evaluation, prompt management and supportive care are essential.

Rather than focusing strictly on a specific time frame, a comprehensive clinical assessment, symptom management, and monitoring of the patient’s response to empirical treatment are crucial. The chosen temperature threshold helps distinguish persistent fever from normal variations, but it can be adjusted based on individual circumstances and evidence availability.

To gain valuable insights into FUO evaluation and management, a proposed study aims to explore the disease profile by considering different temperature ranges and durations of undiagnosed days rather than considering the duration of illness and using a set of obligatory investigations. This research could refine diagnostic criteria, guide clinicians, and improve the timely identification and management of undiagnosed fever cases.

By considering a range of temperature thresholds and durations, the study acknowledges potential variations in clinical practice, healthcare resources, and disease prevalence across regions and patient populations. The findings may shed light on the identification of underlying causes and enhance our understanding of geographic and temporal variations in FUO.

In conclusion, FUO presents a diagnostic challenge, but adapting the criteria based on local healthcare resources allows for a more practical approach. Prompt management and a flexible diagnostic timeline are essential to ensure timely and effective care for patients. The proposed study could provide valuable insights to refine diagnostic criteria based on non-diagnosis rates and different temperature ranges and improve management strategies for undiagnosed fever cases.

## METHODOLOGY

The study was conducted at a tertiary care hospital, All India Institute of Medical Sciences (AIIMS) in Rishikesh, Uttarakhand. It followed a retrospective and prospective Longitudinal-Exploratory study design for a duration of 12 months from May 2022. The study population included patients with fever admitted to AIIMS Rishikesh between January 1, 2018, and December 31, 2022, meeting the inclusion criteria. The sample size was 228 patients, selected through universal sampling. Inclusion criteria consisted of adult patients with documented fever of ≥99.1°F on at least two occasions and fever lasting for more than three days. Exclusion criteria included patients diagnosed with a cause for their fever within three days of hospitalization. Data was collected from five departments of the hospital (General Medicine, Gastroenterology, Neurology, Pulmonary Medicine, Nephrology, and Obstetrics and Gynecology). Retrospective data were obtained from the hospital’s MRD section, and prospective data were collected by following patients who met the inclusion criteria. Data were recorded in an Excel sheet. The study proposed new definitions for FUO based on different ranges of duration of non-diagnosis (3-21 days, >21 days), temperature ranges (99.1°F-100.9°F and ≥101°F) and durations of hospitalization (3-7 days and >7 days) and the frequency of each definition was measured. The statistical analysis involved presenting categorical variables as numbers and percentages, and continuous variables as mean ± SD and median (IQR). The prevalence of FUO and different definitions were calculated using numbers and percentages. A Fisher’s Exact Test was used to compare etiology, and a Chi-Squared Test was used to compare outcomes. A significance level of p<0.05 was considered statistically significant. Data was entered into Microsoft Excel and analyzed using IBM SPSS version 26.0. The study received ethical approval from the Institutional Ethics Committee, AIIMS Rishikesh (No. 43/IEC/PGM/2022), and ensured confidentiality and de-identification of participants’ data. There were no conflicts of interest involving financial interests or organizations.

### PROPOSED DEFINITION

**PROPOSED DEFINITION:**
| DEFINITION | DAYS OF<br>UNDIAGNOSIS | TEMPERATURE | DAYS OF<br>HOSPITALIZATION |
| --- | --- | --- | --- |
| A | 3-21 | 99.1F-100.9F | 3-7 |
| B | 3-21 | 99.1F-100.9F | >7 |
| C | 3-21 | >/= 101F | 3-7 |
| D | 3-21 | >/= 101F | >7 |
| E | >21 | 99.1F-100.9F | 3-7 |
| F | >21 | 99.1F-100.9F | >7 |
| G | >21 | >/= 101F | 3-7 |
| H | >21 | >/= 101F | >7 |

## RESULTS

This study aimed to estimate the prevalence of Fever of Unknown Origin (FUO) based on different proposed definitions and analyze the etiology as well as outcomes of FUO patients.

After considering inclusion as well as exclusion criteria, 228 patients were included in the study. The distribution of participants revealed that Definition B (40.8%) is having the highest number of patients followed by Definition A (21.5%), followed by Definition D (19.3%), followed by Definition C (12.3%), While only 5 patients met the criteria for the classical definition of FUO [Definition H] (in terms of temperature and duration of hospitalization). By closely observing the patients in Definition B group, around 36.5% patient remained non-diagnosed between >7 to 10 days which is highest among the group. This data indicates that around 62% of the patients remained undiagnosed before 21 days of hospitalization and started on treatment and they are having a temperature range between 99.1F to 100.9 F. Among the rest 38% of patients, 31.6% of patients remained undiagnosed and started on treatment within 21 days and they were having temperature values of >/=101F. Thus the majority of patients (94%) underwent diagnostic workups and started treatment within 21 days of hospitalization. When comparing the different definition groups, Definition B had the highest number of cases attributed to infection (87.1), malignancy, and autoimmune diseases. There was no significant difference in the distribution of infection, malignancy, and autoimmune etiology among the various definition groups.

There was no significant difference in the outcome of these patients at discharge among these definitions. Although only 129 patients could be followed up for outcomes at 180 days, there was no significant difference between the outcome at 180 days follow-up. Infection was the most common etiology among all the proposed definitions followed by autoimmune disorder and malignancy. This etiological trend is similar to the majority of FUO studies done in the non-western world. While till yet, no study has been done considering different duration and temperature criteria, it is not feasible to compare these findings.

### Distribution of the Participants in Terms of ‘Definition’

**Distribution of the Participants in Terms of 'Definition'**
| **Definition** | **Frequency** | **Percentage** | **95% CI** |
| --- | --- | --- | --- |
| A | 49 | 21.5% | 16.5% - 27.5% |
| B | 93 | 40.8% | 34.4% - 47.5% |
| C | 28 | 12.3% | 8.5% - 17.4% |
| D | 44 | 19.3% | 14.5% - 25.1% |
| E | 1 | 0.4% | 0.0% - 2.8% |
| F | 7 | 3.1% | 1.4% - 6.5% |
| G | 1 | 0.4% | 0.0% - 2.8% |
| H | 5 | 2.2% | 0.8% - 5.3% |

### Time to diagnose among Definition B

**Time to diagnose among Definition B:**
| **Time to Diagnose (Days)** | **Frequency** | **Percentage** |
| --- | --- | --- |
| 3-7 days | 14 | 15% |
| >7- 10 days | 34 | 36.5% |
| >10- 14 days | 26 | 28% |
| >14-21 days | 19 | 20.5% |

### Association Between Definition and Outcome

| Outcome | Definition |  |  |  |  |  |  |  |  | Chi-Squared Test |  |
| --- | --- | --- | --- | --- | --- | --- | --- | --- | --- | --- | --- |
| | A | B | C | D | E | F | G | H | Total | $\chi^2$ | P Value |
| Alive | 37<br>(75.5%) | 83<br>(89.2%) | 23<br>(82.1%) | 40<br>(90.9%) | 0<br>(0.0%) | 7<br>(100.0%) | 1<br>(100.0%) | 5<br>(100.0%) | 196<br>(86.0%) | 27.937 | 0.142 |
| Unstable Vital | 4<br>(8.2%) | 3<br>(3.2%) | 1<br>(3.6%) | 1<br>(2.3%) | 0<br>(0.0%) | 0<br>(0.0%) | 0<br>(0.0%) | 0<br>(0.0%) | 9<br>(3.9%) |  |  |
| Poor Prognosis | 0<br>(0.0%) | 2<br>(2.2%) | 0<br>(0.0%) | 2<br>(4.5%) | 0<br>(0.0%) | 0<br>(0.0%) | 0<br>(0.0%) | 0<br>(0.0%) | 4<br>(1.8%) |  |  |
| Death | 8<br>(16.3%) | 5<br>(5.4%) | 4<br>(14.3%) | 1<br>(2.3%) | 1<br>(100%) | 0<br>(0.0%) | 0<br>(0.0%) | 0<br>(0.0%) | 19<br>(8.3%) |  |  |
| Total | 49<br>(100.0%) | 93<br>(100.0%) | 28<br>(100.0%) | 44<br>(100.0%) | 1<br>(100.0%) | 7<br>(100.0%) | 1<br>(100.0%) | 5<br>(100.0%) | 228<br>(100.0%) |  |  |

### Outcome on Follow Up at 180 days

| Outcome on Follow Up | Definition |  |  |  |  |  |  | Fisher's Exact Test |  |
| --- | --- | --- | --- | --- | --- | --- | --- | --- | --- |
| | A | B | C | D | F | H | Total | $\chi^2$ | P Value |
| Alive | 25<br>(83.3%) | 35<br>(74.5%) | 17<br>(94.4%) | 21<br>(75.0%) | 3<br>(100.0%) | 2<br>(66.7%) | 103<br>(79.8%) | 4.945 | 0.393 |
| Death | 5<br>(16.7%) | 12<br>(25.5%) | 1<br>(5.6%) | 7<br>(25.0%) | 0<br>(0.0%) | 1<br>(33.3%) | 26<br>(20.2%) |  |  |
| Total | 30<br>(100.0%) | 47<br>(100.0%) | 18<br>(100.0%) | 28<br>(100.0%) | 3<br>(100.0%) | 3<br>(100.0%) | 129<br>(100.0%) |  |  |

## DISCUSSION

The study on Fever of Unknown Origin (FUO) in a tertiary care hospital in North India provides valuable insights into the prevalence, etiology, and outcomes of FUO patients based on different proposed definitions. The findings shed light on the challenges faced in diagnosing and managing FUO cases in a resource-constrained setting, and they highlight the need for revising the current criteria used for defining FUO.

The study included 228 patients who met the inclusion and exclusion criteria, and their distribution among different proposed definitions revealed that Definition B was the most common (40.8%), followed by Definitions A, D, and C. Surprisingly, only a small number of patients (5 patients, 2.2%) met the classical definition of FUO (Definition H) in terms of temperature and days of hospitalization. This finding raises concerns about the applicability of the classical definition.

The data indicates that a significant proportion (62%) of patients remained undiagnosed before 21 days of hospitalization and were started on treatment. Notably, these patients had a temperature range between 99.1°F to 100.9°F, which is below the classical cutoff of >101°F for defining FUO. On the other hand, 38% of patients had temperatures of ≥101°F and remained undiagnosed before starting treatment. This suggests that using a single temperature cutoff for all patients may not be suitable, and a more nuanced approach is needed.

The majority of patients (94%) underwent diagnostic workups and started treatment within 21 days of hospitalization, regardless of the definition used. This highlights the importance of early evaluation and treatment for FUO patients, especially in resource-constrained settings like India, where access to advanced laboratory facilities may be limited.

Comparing the different definition groups, Definition B was associated with the highest number of cases attributed to infection, malignancy, and autoimmune diseases. However, there was no significant difference in the distribution of these etiologies among the various definition groups. This indicates that the choice of definition did not significantly impact the distribution of underlying causes of FUO, suggesting that all proposed definitions captured similar patterns of etiological trends.

Another critical finding is that there was no significant difference in patient mortality at discharge and the 180-day follow-up among the various definitions. This suggests that the choice of definition did not impact the short-term and medium-term prognosis of FUO patients.

It further supports the notion that revising the criteria to better suit the local patient population’s needs and resource constraints would not compromise patient outcomes.

Infection was found to be the most common etiology among all the proposed definitions, followed by autoimmune disorders and malignancy. This trend is consistent with other FUO studies conducted in non-western regions, highlighting the importance of considering regional epidemiology when defining FUO criteria.

The study’s results underscore the need for a revised approach to defining FUO. Due to limitations in laboratory facilities and socioeconomic factors affecting patient care, modifying the FUO criteria to allow for earlier diagnosis (considering the days of non-diagnosis rather than illness duration of >3 weeks) and considering a lower temperature cutoff (>99.1°F) may be more appropriate for this patient population. Such modifications would enable clinicians to initiate empirical treatment based on clinical judgment in a timely manner.

## CONCLUSION

The study provides valuable evidence suggesting that the criteria for diagnosing FUO need revision. Adapting the criteria to better reflect the local patient population’s needs and resource constraints could improve the early diagnosis and management of FUO cases, ultimately leading to better patient outcomes. Further research and collaboration within the medical community are essential to developing comprehensive and context-specific guidelines for diagnosing and managing FUO.

## Data Availability

It will be available by contacting the corresponding author.

## Contributors

PD contributed to the data collection, data analysis, and was involved in manuscript writing.

RC involved in data analysis and manuscript writing

PKP gave the concept, interpreted analysis, critically reviewed the draft, and approved it for publication along with all authors.

RK, RG, RD, AT, SS, KK reviewed the draft.

## Data sharing

It will be made available to others as required upon requesting the corresponding author.

## Acknowledgment

None

